# Emergence of COVID-19 Variant XFG not linked to Higher Rates of Sore Throat or altered severity: Evidence from participatory surveillance in The Netherlands

**DOI:** 10.1101/2025.10.15.25338082

**Authors:** Gesa Carstens, Eva Kozanli, Wanda Han, Marie Gruters, Rianne van Gageldonk, Susan van den Hof, Albert Jan van Hoek, Dirk Eggink

## Abstract

Since the emergence of SARS-CoV-2, the virus causing COVID-19, it evolved into different lineages and sub-variants. New genetic variants of the virus can result in a different transmissibility, immune evasion, as well as different symptoms and severity of disease. When new variants appear, media report often anecdotal observations of unique or differentiating symptomatology of these new variants. Here, we investigate reported elevated levels of sore throat linked to the new variant XFG using an evidence-based method.

## Emergence and impact of novel variants

After the ancestral wild-type variant, Alpha and Delta, a wide variety of Omicron variants have circulated. Initial variants like Alpha and Delta were mostly associated with increased transmissibility (1), while the rise and spread of Omicron variants since late 2021 was mostly associated with antigenic changes (2). Both altered transmissibility and antigenic evolution can change the epidemiology. Furthermore, these genetic changes can affect symptomatology or severity, not necessarily leading to more infections but leading to an altered or increased disease burden. Since the rise of the Omicron, SARS-CoV-2 infections have been reported to be milder(3, 4) compared to Delta with relatively more upper respiratory and common cold-like symptoms (sore throat, congestion, cough, fatigue), instead of lower respiratory symptoms. In addition, loss of taste or smell has become less common compared to the early days of the COVID-19 pandemic (5).

In the summer of 2025, media reports appeared in (digital) newspapers, weblogs, and social media about altered manifestation of COVID-19, possibly linked to the novel SARS-CoV-2 Omicron variant XFG (on social media often named by its unscientific name ‘Stratus’) compared to the previously dominant Omicron variants JN.1 and KP.3; with a particular focus on sore throat and hoarse voice(6-8). However proper scientific basis for such statements are rare to non-existent.

The participatory community surveillance study *Infectieradar (9)* in The Netherlands has been analyzing respiratory symptoms and causative respiratory pathogens since 2020. Participants (39,937) weekly report the presence of respiratory symptoms and a corresponding health score (a scale from 0 to 100, with 100 indicating the best possible health and 0 indicating the worst possible health). In addition, they are asked to perform a COVID-19 self-test when reporting respiratory symptoms and a representative sub-selection of participants with respiratory symptoms is asked to send in a nose-throat swab to the National Institute for Public Health and the Environment (RIVM) for molecular diagnostics and whole genome sequencing of detected SARS-CoV-2. This allows detailed comparison of symptomatology, including sore throat, of different epidemic waves and variants over time. Hoarse voice is not a symptom in our standard questionnaire, and unlisted symptoms are inconsistently reported, hence our analysis focusses on sore throat.

## COVID-19 Epidemiology 2024 vs 2025

Epidemiological trends revealed a distinct pattern in SARS-CoV-2 positive cases between the waves in 2024 and 2025. Cases started to rapidly increase in 2024 in week 22, peaking around week 29 and followed by a subsequent decline and new rise in incidence around week 40, together considered as the 2024 wave in further analyses and descriptions. 2025 was characterized by a lower, yet steadily increasing number of cases starting week 30 with a more rapid surge as of week 37 (Figures 1A and 1B). According to the Dutch national SARS-CoV-2 genomic surveillance (10), which also includes sequences from the Infectieradar nose-throat swab, the JN.1 subvariant KP.3 was predominant during the 2024 wave, accounting for the majority of infections (60-79%) each week (Figure 1C). By contrast, in 2025, the XFG variant quickly rose in dominance, rapidly increasing from a prevalence of approximately 50% to over 90% of all infections by week 38 (Figure 1D). To allow comparison of symptoms and severity during the same time of year, and investigate whether symptoms are unique, indicative, or differentiating for XFG, we compared COVID-19 infections from *Infectieradar* during the emergence of XFG in 2025 (week 28-40; n=1,539), with infections from the same period in 2024, representative for the previous dominant variant KP.3 (n=605). Both sub-cohorts displayed similar background characteristics (Table 1).

**Table 1:** Participant characteristics.

|  | **2024** | **2025** |
| --- | --- | --- |
|  | **N = 1,539** | **N = 605** |
| **Sore throat** | 1,096 (71%) | 448 (74%) |
| **Sex** |  |  |
| female | 1,018 (66%) | 416 (69%) |
| male | 521 (34%) | 189 (31%) |
| **Age group (years)** |  |  |
| <25 | 21 (1.4%) | 7 (1.2%) |
| 25-39 | 172 (11%) | 73 (12%) |
| 40-49 | 265 (17%) | 119 (20%) |
| 50-64 | 642 (42%) | 232 (38%) |
| 65+ | 439 (29%) | 174 (29%) |
| **Education level** |  |  |
| lower/none | 13 (0.9%) | 2 (0.3%) |
| middle | 529 (35%) | 203 (34%) |
| higher | 981 (64%) | 396 (66%) |
| **Smoking** | 106 (6.9%) | 40 (6.6%) |
| **Allergy** | 603 (39%) | 246 (41%) |
| **Asthma** | 138 (9.0%) | 74 (12%) |
| **Diabetes** | 53 (3.4%) | 15 (2.5%) |
| **Chronic lung disease** | 38 (2.5%) | 16 (2.6%) |
| **Cardiovascular disease** | 156 (10%) | 56 (9.3%) |

**Figure 1:**
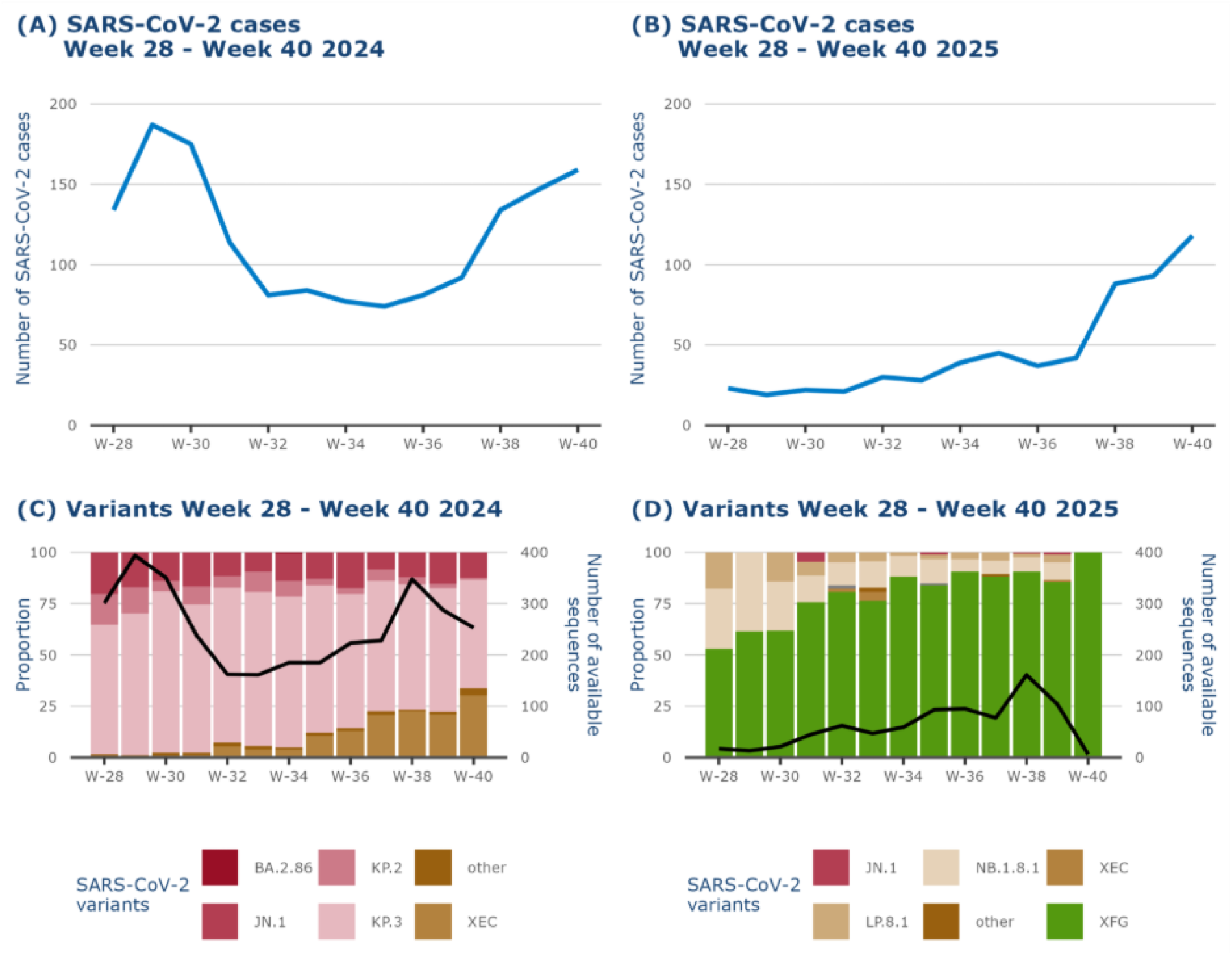
Overview of SARS-CoV-2 positive cases in Infectieradar and variant distribution in the Netherlands. SARS-CoV-2 positive cases (based on self-test or PCR test) in the community participatory surveillance study Infectieradar during week 28–40 2024 (A) and week 28-40 in 2025 (B). Distribution of SARS-CoV-2 variants in the Netherlands, as monitored by the national SARS-CoV-2 genomic surveillance program (kiemsurveillance(10)), during the same periods in 2024 (C) and 2025 (D). Variant data include sequenced samples from a random selection of symptomatic Infectieradar participants, representing variant circulation within this population. The black lines indicate the number of available sequences per week.

## Shifting COVID-19 symptomatology and severity over time

To compare symptomatology, we performed a multivariable logistic regression to estimate the association between year of infection and the presence of specific symptoms, adjusting for age group, sex, and comorbidity status. A running nose, cough, sneezing and a sore throat were the most reported symptoms during both epidemiological COVID-19 waves. When focusing on sore throat we observed that although a sore throat was reported slightly more often in 2025, it was already high in 2024: 74% vs 71% in 2024. The difference was not significant in our multivariate logistic analysis (Figure 2A; OR: 1.14, 0.92-1.42 95% CI). Furthermore, sore throat was not uniquely elevated, as many other symptoms increased slightly, however none of these were significantly elevated neither. “Hoarse voice” (and all relevant synonyms) was only rarely reported in our open field of the questionnaire: it was only mentioned two times in 2024 and two times in 2025, suggesting it is a not considered an important symptom by the *Infectieradar* participants during either COVID-19 wave.

**Figure 2:**
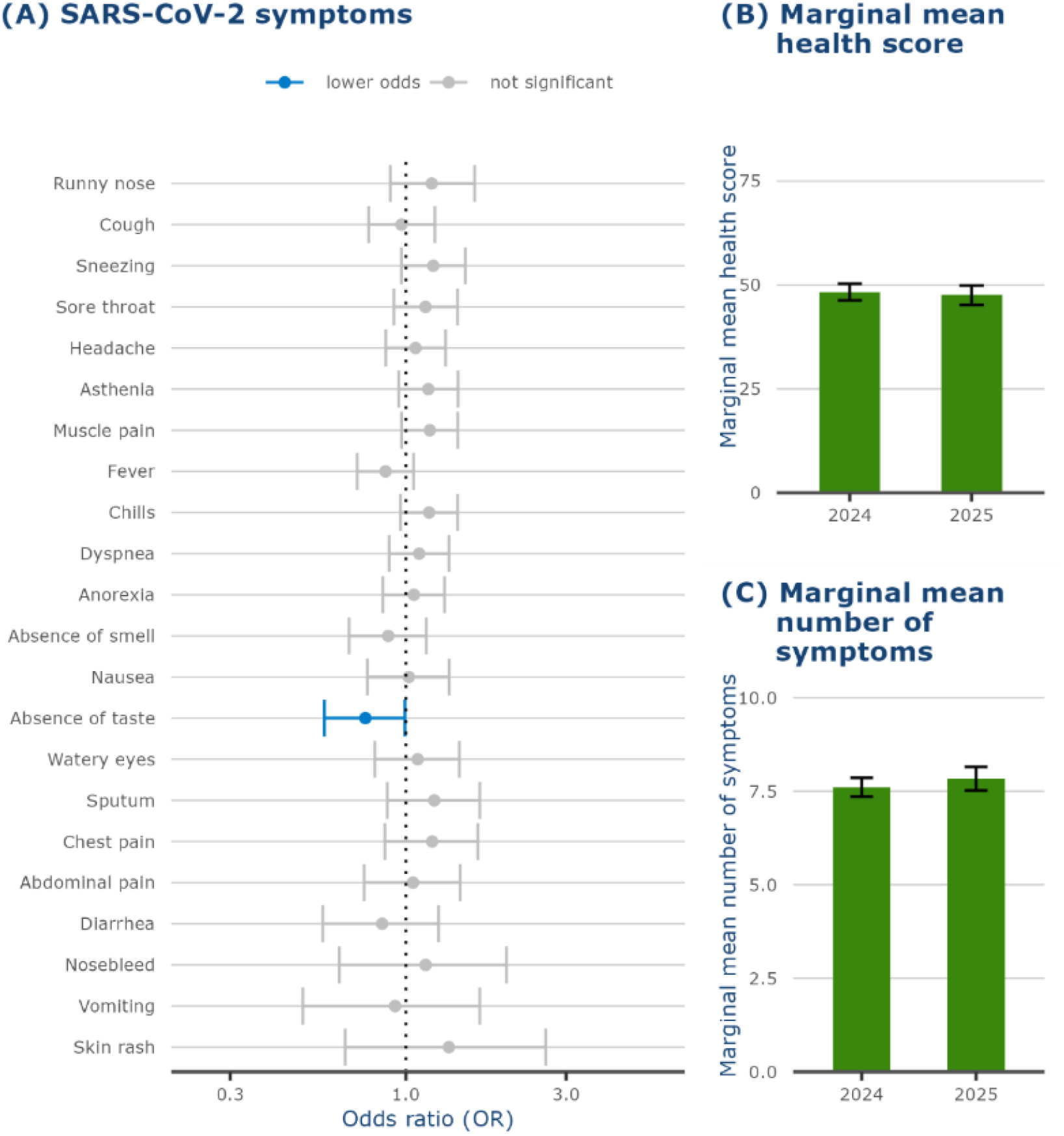
Comparison of COVID-19 symptomatology in Infectieradar participants, 2024 vs. 2025. Symptom reporting of SARS-CoV-2 positive Infectieradar participants (confirmed by self-test or PCR) during weeks 28-40 2024 (n = 1,539) and 2025 (n = 605), representative of KP.3 and XFG infections respectively, were compared using multivariate regression models corrected for age group, sex and presence of comorbidities using 2024 as reference group (A). The lowest self-reported health score during infection was compared for the same periods using marginal means corrected for age group, sex and presence of comorbidities (B). The number of symptoms reported during SARS-CoV-2 infection was compared for the same periods using marginal means corrected for age group, sex and presence of comorbidities (C).

The only significant change between the two periods was a further decline in presence of loss of taste compared to 2024 from 17% to 13% (OR: 0.76, 0.57-0.99 95% CI; Figure 2A). This observation aligns with the general observation that loss of smell and tase is less present during COVID-19 in recent years, either related to novel Omicron variants with altered tropism or the possible role of pre-existing immunity(11).

Although the prevalence of sore throat is not altered, the symptom might present more apparent, resulting in experiencing more severe disease. To assess possible difference in disease severity, we investigated the impact of SARS-CoV-2 infection during both COVID-19 waves, on health status using a linear regression model, evaluating the association between year of infection and the lowest reported health score during illness. This analysis controlled for age, sex, comorbidities (asthma, diabetes, chronic lung disease, cardiovascular disease, chronic kidney disease and immunodeficiency), and the non-linear effect of days since symptom onset. In addition, we compared symptom burden using a multivariable Poisson regression model, with the total number of symptoms as the dependent variable and year of infection, age group, sex, and comorbidity status as independent variables. Estimated marginal means (EMMs) indicated that the lowest health score during infection was similar between the two years. On top, the adjusted mean number of symptoms reported was almost identical (7.6 in 2024 and 7.8 in 2025). Therefore, we can conclude that the overall severity experienced during the two COVID-19 waves was similar and there is no evidence for altered disease because of more severe sore throat.

## Discussion / Conclusion

Our community participatory surveillance uniquely investigates respiratory infections and burden of disease and tracks emerging trends in real-time. We here describe the COVID-19 symptoms in the general population during epidemiological waves in 2024 and 2025, representing infections with SARS-CoV-2 variants KP.3 and XFG respectively. In our data, XFG infection is not significantly associated with higher prevalence of sore throat, nor does it appear to present unique symptomology compared to previous SARS-CoV-2 variants such as KP.3 as suggested by on-line media in late summer and fall of 2025. While slightly elevated trends for various symptoms were observed, these were not significant. The prevalence of loss of taste and smell among those infected with SARS-CoV-2 continued to decline. We were not able to draw conclusions about the media reports on a higher prevalence of a hoarse voice, as these were not part of the standard symptom list in our questionnaire. However, these were not frequently reported as an additional symptom, indicative that these are not considered an important symptom by the *Infectieradar* participants. Overall, Infectieradar is able to monitor possible altered symptoms and severity during COVID-19 in the general public in real time, showing symptomology and disease severity during XFG infection is comparable to earlier variants observed in the 2024 wave.

## Data Availability

The processed data required to reproduce the findings of this study are available from the corresponding author upon reasonable request. Due to confidentiality concerns, access to the raw data may be restricted.

